# Regularized COVID-19 Forecast Ensemble Methods

**DOI:** 10.1101/2023.05.12.23289872

**Authors:** Alexandra Stephens, Luke C. Mullany, Matt Kinsey, Paul Nicholas, Jeffrey Freeman, Kaitlin Rainwater-Lovett

## Abstract

Forecasts of COVID-19 outcomes play an essential role in alerting public health and government officials to the trajectory of the pandemic. The sudden and critical need for these forecasts spurred both the proliferation of diverse epidemiological transmission models from academia and industry across the United States and efforts to standardize and curate these model outputs. In many scientific domains, ensemble models, where individual forecasts are aggregated into one, have demonstrated smaller forecasting error than the individual models from which they are constructed. Using COVID-19 deaths as an index outcome, we developed and evaluated several ensemble approaches where point forecast models were combined via weighted sums based on historical individual model or ensemble model performance. We found that a simple method that minimized the error of the past performance of individual models and used L2 regularization to encourage broader distribution of weights across models outperformed a baseline mean ensemble and all other tested methods across US states for both absolute error and weighted interval scores. This suggests that performance-based ensembles can produce accurate forecasts despite training on only point forecasts and recent historical data, provided that sufficient regularization and constraints are used to capture uncertainty. Availability of an accurate and explainable ensemble forecast model can increase trust among stakeholders and the general public, thus bettering preparedness and response efforts during the COVID-19 pandemic.

## 1 Introduction

The COVID-19 pandemic created an immediate demand for timely and accurate forecasts of cases, deaths, and hospitalizations. Starting in March of 2020, the COVID-19 Forecast Hub (“the Hub”) began collecting weekly COVID-19 forecasts in a standardized format (covid19forecasthub.org). Up to 70 research teams have submitted forecasts to the Hub, creating a diverse and ever-growing historical record of real-time COVID-19 forecasts. These individual, or component, models are submitted in the form of single point forecasts and multiple quantile forecasts to provide uncertainty bounds. Given the large quantity of models and wide ranges of uncertainty within each model, the spectrum of probable outcomes becomes too wide for stakeholders to utilize to make informed decisions. Accurate forecasts are also essential to help governments and health departments prepare for and respond to public health crises. Additionally, forecast reversals undermine public trust in COVID-19 forecast models (6). To more adequately deliver trustworthy and actionable model results, a single and robust forecast model with reliable uncertainty is required.

When choosing a final model for any predictive modeling or machine learning application, cross-validation is often used to compare various models and parameters where the model-parameter combination with the smallest prediction error on the test data is selected. However, combining multiple predictive models into an *ensemble* has long been known to often out-perform the individual models chosen through cross-validation from the same set (2). This concept has been applied to forecasting, where even simple average ensembles show steady performance in influenza forecasting applications (8). For COVID-19, the Hub’s established ensemble model has been producing weekly forecasts by, at first, taking an average across the subset of models with one-to four-week-ahead forecasts, and, later in July 2020, switching to using a median of the same subset. This ensemble has displayed the best overall probabilistic accuracy compared to the component models, and the authors emphasize the importance of combining forecast models (4), (11). Neither the mean nor the median take model characteristics, such as individual or combined historical performance, into account. There is much evidence to suggest that combining forecast models unequally as a weighted-sum ensemble may provide even more accurate forecasts than an equal-weight ensemble (7), (12), (14).

Linearly combining predictive models, such as regressions, through minimizing the error of combined model predictions is referred to as *stacking* (17). The stacked algorithm is meant to learn the relationship between a combination of models and the observed data (14). Stacking is one of many forms of ensemble development and it has been successful in its application to regression and classification models, where the weights are constrained to be non-negative (2), (17). This algorithm does not necessarily reward individual model performance, but seeks the optimal combination of models that are preferably very different from one another (2). Ting and Witten (15) noted issues in this framework for classification and suggested combining model confidence instead of single valued predictions.

For disease and weather forecasting applications, ensembles have been created using Bayesian Model Averaging (BMA) and other similar schemes that account for model uncertainty. The resulting weights are a reflection of individual forecast skill (10). The ensemble is framed as a mixture model of probability densities where each observation is believed to have been generated from a single model. Weights are constrained to sum to one to create a probability distribution. To maximize the log-likelihood of this mixture model ensemble, one places weight on models that display the best past performance while accounting for the uncertainty provided by their reported distributions (12). Under a similar probabilistic framework, McAndrew and Reich (7) explored the forecasting abilities of an adaptive ensemble, where a new set of weights were calculated for each forecasting week of an influenza season and the model was trained on only the current season’s historical data. Regularization of weights given a uniform Dirichlet prior was found to be advantageous in this adaptive setting where data was prone to revision.

While the probabilistic framework has shown success in influenza and other forecasting applications (12), the mathematical complexity of this approach might be at odds with the need to provide readily accessible results to a broad range of stakeholders, including the general public. Motivated to find a simple and robust ensemble method, we developed and evaluated several optimization methods on COVID-19 point forecasts with incident deaths as the target outcome. Death was selected as a model outcome because deaths were better captured than cases during the pandemic. The uncertainty surrounding case estimation was due to fluctuating SARS-CoV-2 testing volumes and often unreported asymptomatic infections (9), (13). We implemented two ensemble frameworks: optimization of the performance of the combined models (as is traditionally done in stacking, constrained optimization, and linear regression), and combining models based on individual performance, inspired by the intuitive understanding of BMA. All ensemble weights were constrained to be non-negative and sum to one, as is common in the stacking ensemble literature. We added regularization terms and constraints that encouraged a broader distribution of weights across models, similar to the regularization described in the previous paragraph which was integrated via an equal-weight prior distribution (7). We describe a novel method that appears to provide forecasts with smaller absolute error and weighted interval scores than the baseline and other ensemble methods, and provide an overview of the many advantages of this simple yet effective methodology compared to previous work. We compare and contrast our approaches and comment on the challenges of COVID-19 ensemble forecasting.

## 2 Methods

### 2.1 Data

Component model forecasts of 1-week-ahead incident deaths were obtained from the Hub and used for training and testing ensemble methods. Forecast location targets included the United States (US), all 50 states, and the District of Columbia (DC). The observed data were derived from the Johns Hopkins University Center for Systems Science and Engineering COVID-19 Dashboard (5). A rolling 7-day sum was applied to the daily empirical data to match the weekly incident death model forecasts submitted to the Hub. The training data were further smoothed by fitting a natural cubic spline Poisson regression model to the weekly incident deaths in each training period, and the full time series of 7-day sums was similarly smoothed to create the testing data.

To create a singe set of ensemble weights, training data consisted of 15 1-week-ahead predictions from *M* valid models for a particular location, as well as the weekly observed incident deaths for the same 15 weeks and location. The data were used as inputs for an ensemble method, and the resulting weights were applied to the following week’s forecasts provided by the same *M* models to create an out-of-sample 1-week-ahead ensemble forecast. This constitutes a single “test” data point, thus this process is repeated as a sliding time window to create multiple weeks of out-of-sample testing ensemble forecast data.

Inclusion criteria were applied to each training and testing dataset to remove certain models before ensemble weight optimization. The criteria for component model point and quantile forecasts was decided in conjunction with the 15-week training time window. The Hub required incident death forecasts were provided as a point forecast along with 23 quantile forecasts ranging from 0.01 to 0.99, thus models must have had a point and all 23 quantile forecasts on the test date. Additionally, models must have had no more than 20% missing point estimates during the training period. We selected the <20% threshold to balance the benefit of including more models (i.e features) during optimization with the cost of training on incomplete and possibly misleading representations of models. Similarly, the 15 week time window helped to keep this balance: the longer the time window, the more likely models are to be missing a portion of data, but too short a time window could lead to overfitting. Additional time windows and thresholds were examined during preliminary analyses; a full parametric analysis could be completed in future work.

### 2.2 Ensemble Methods

Two distinct ensemble methods referred to as *Combined Error (CbEr)* and *Individual Error (IndEr)* were developed and applied. The CbEr methods minimized the error of the weighted sum of models, while the IndEr methods minimized the weighted sum of the individual model error. Regularization and constraints were implemented for both groups of methods to encourage smaller, more distributed weights. For comparison, two baseline models were implemented. The final (non-baseline) methods each involve a global parameter, thus a hyperparameter selection was performed for all methods by examining 15 weeks of out-of-sample ensemble forecasts.

For each ensemble method, location, and set of training dates, a set of weights *w*_*j*_ were optimized in order to minimize an objective subject to constraints. The weights were constrained to be nonzero and sum to 1, creating a weighted sum of component models. Consider the set of *x*_*i,j*_ point forecasts for a particular location, where *i* indexes the forecast date, *i* = 1, 2, …*T*, and *j* indexes the set of valid models, *j* = 1, 2, …*M* . The ensemble point forecast for the test date *i, z*_*i*_, is defined as follows:

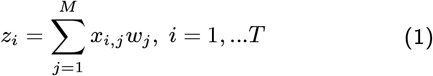

Given that each component model produced 23 quantile forecasts for the incident death target, let *x*_*i,j,q*_ where *q* = 1, 2, …23 represent the component model forecast for quantile *q* and *z*_*i,q*_ represent the ensemble forecast for quantile *q*. Let the absence of a *q* indicate *x*_*i,j*_ and *z*_*i*_ are point forecasts. For ensemble evaluation, quantile forecasts were calculated as a weighted sum, where the same weight is applied to each quantile *q* for date *i*,

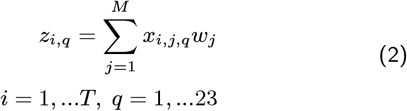

All optimizations were completed using the Python library SciPy’s minimize function with the L-BFGS-B algorithm (Limited-memory BFGS), a bounded quasi-Newton optimization method (3), (16). The weights were bounded to be between 0 and 1 and were rescaled to sum to 1 within each iteration.

A *Baseline Mean* method was evaluated alongside the optimization methods as a means of comparison. The baseline is an equal-weight sum of all valid models, 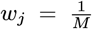 This method is common in the infectious disease ensemble literature and appears as both a standalone method and a baseline for comparison against unequal weighted methods (7), (11), (12).

### 2.2.1 Combined Error (CbEr) Ensemble Methods

The loss function to be minimized for these methods is the root-mean-square error (RMSE),

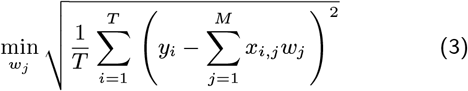

where the weights *w*_*j*_ *∀ j* minimize the RMSE of the weighted sum ensemble, and the smoothed observed data are represented as *y*_*i*_. This optimization with no additional constraints or regularization terms was also included in the set of evaluated methods, and will be referred to as the *CbEr Baseline*. We introduce two variations of this optimization:

*a. Combined Error with L2 regularization (CbEr-L2)*: L2 regularization, 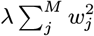, is added to the base optimization in Equation 3 where *λ* is the regularization coefficient decided during hyperparameter selection. This formulation is similar to a constrained Ridge Regression, but with RMSE instead of the sum of squares.

*b. Combined Error with Lower Bound (CbEr-LB)*: weights are first optimized as in Equation 3, then a portion of the weights are redistributed to the models with weight less than a specified lower bound such that *w*_*j*_ *≥ LB ∀j*.

### 2.2.2 Individual Error (IndEr) Ensemble Methods

Here, we minimize a weighted sum of the RMSE of each individual model:

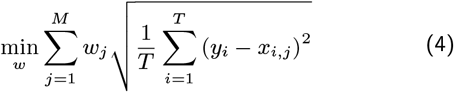

We note that if this equation is optimized directly with no regularization and only the basic constraints on the *w*_*j*_, the single model with the smallest RMSE will receive a weight of 1, while all other models will have zero weight.

To encourage the inclusion of models, two variations of this optimization were implemented:

a. *Individual Err with L2-Regularization (IndEr-L2)*: L2 regularization, 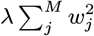 is added to the base objective function in Equation 4. This is similar to 2.2.1 (a), with the difference in objective function formulation.

b. *Top-N Individual Error (IndEr-TN)*: non-zero weights are constrained to be equal, and the number of non-zero weights must be no more than *N*. Simply: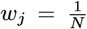 for the *N* models with the smallest RMSE.

### 2.2.3 Hyperparameter Selection

Each ensemble method required the selection of a hyperparameter before optimizing for ensemble weights: the coefficient *λ* in L2 regularization, the value of the lower bound *LB* in CbEr-Lb, and the value of *N* in IndEr-TN. For the first two parameters, there is slightly more nuance than selecting a constant value. For regularization, we set 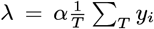 to scale the term appropriately against Equations 3 and 4, leaving us to find a value for *α*. If *α* = 1, the term would be equal to the average *y*_*i*_ empirical values. If *α* = 0, we are simply optimizing Equations 3 and 4. For the lower bound, *LB* must be a function of the total valid models *M*, 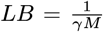 where *γ ≥* 1. If *γ* = 1, we have an equal-weight sum.

To select hyperparameter values, 15 weeks of out-of-sample forecasts were generated with each ensemble method for a spectrum of *α, γ*, and *N* values. This was completed for the US and a few select states with larger populations from mid December 2020 through late March 2021. An example of what may have been considered during this process is shown in Figure 1, where the *α* parameter is iterated from 0 to .6, and the mean absolute error is calculated on many weeks of ensemble forecasts with each value of *α*. Hyperparameters that roughly minimized the error were selected: the value *α* = 0.3 for both IndEr-L2 and CbEr-L2, N=10 for IndEr-TN, and *γ* = 1.5 where 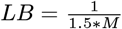for CbEr-Lb.

**Figure 1.**
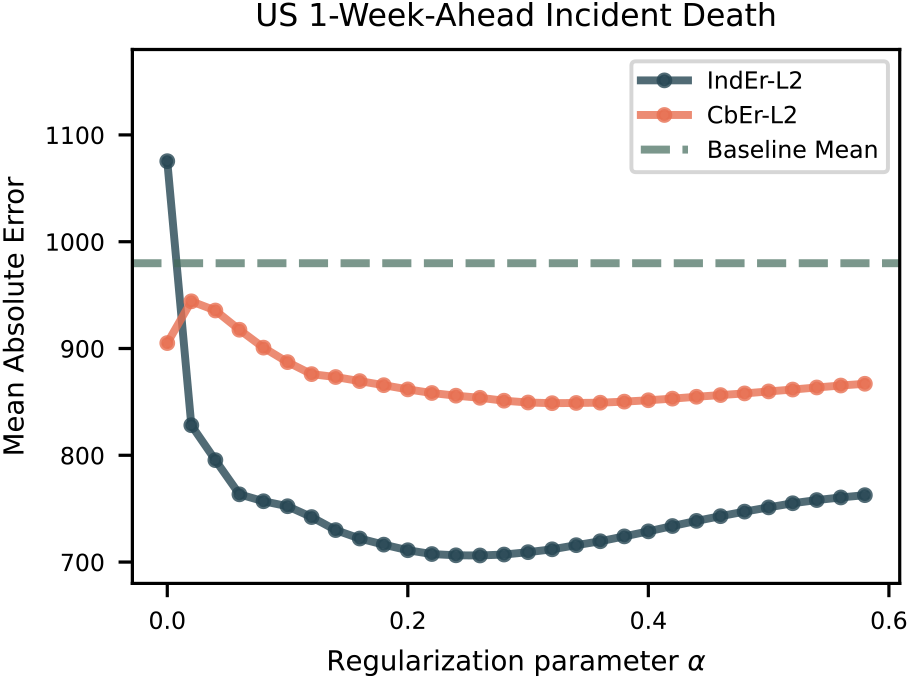
Comparison of the performance of ensembles with L2 regularization across hyperparameter values (*α*) for the 30-week out-of-sample ensemble forecasts test set (August 2020 -March 2021) and smoothed US 1-week-ahead incident deaths. When *α* = 0, the IndEr-L2 method can be interpreted as a selection of the single model with the smallest RMSE during the training period. Weights approach equality as *α* is increased.

## 3 Results

The test dataset comprised 30 weeks of 1-week-ahead model forecasts and smoothed observed weekly incident deaths as described in Section 2.1 beginning in August 2020 and ending in March 2021 for all 50 states and DC. The ensemble forecasts were created using a sliding 15-week training window ending the week before each test date, thus the training data included 45 weeks overall. The absolute errors of the point forecasts and the weighted interval scores (WIS) of the quantile forecasts for each ensemble method, date, and location target were calculated. The weighted interval score is a metric that evaluates forecasts in an interval format and has become widely used for COVID-19 forecasts (1). Next, aggregate statistics on the 30 dates for each method and location were derived, including the mean (resulting in the Mean Absolute Error or MAE and the mean WIS), median, maximum, and standard deviation. The six ensemble methods (i.e., two baseline approaches and four variations) were then ranked for each of the summary metrics within each state, and the average of these 51 ranks are shown in Figure 2. Additionally, the average of the absolute error metrics are given in Table 1. The absolute error is closely aligned with the metric used during optimization, RMSE, while the weighted interval score indicates the resulting ensemble’s ability to predict reasonable uncertainty intervals. Thus, the absolute error metrics provide a more fair comparison of ensemble performance, and the weighted interval score results demonstrate performance of the interval ensemble forecasts.

**Figure 2.**
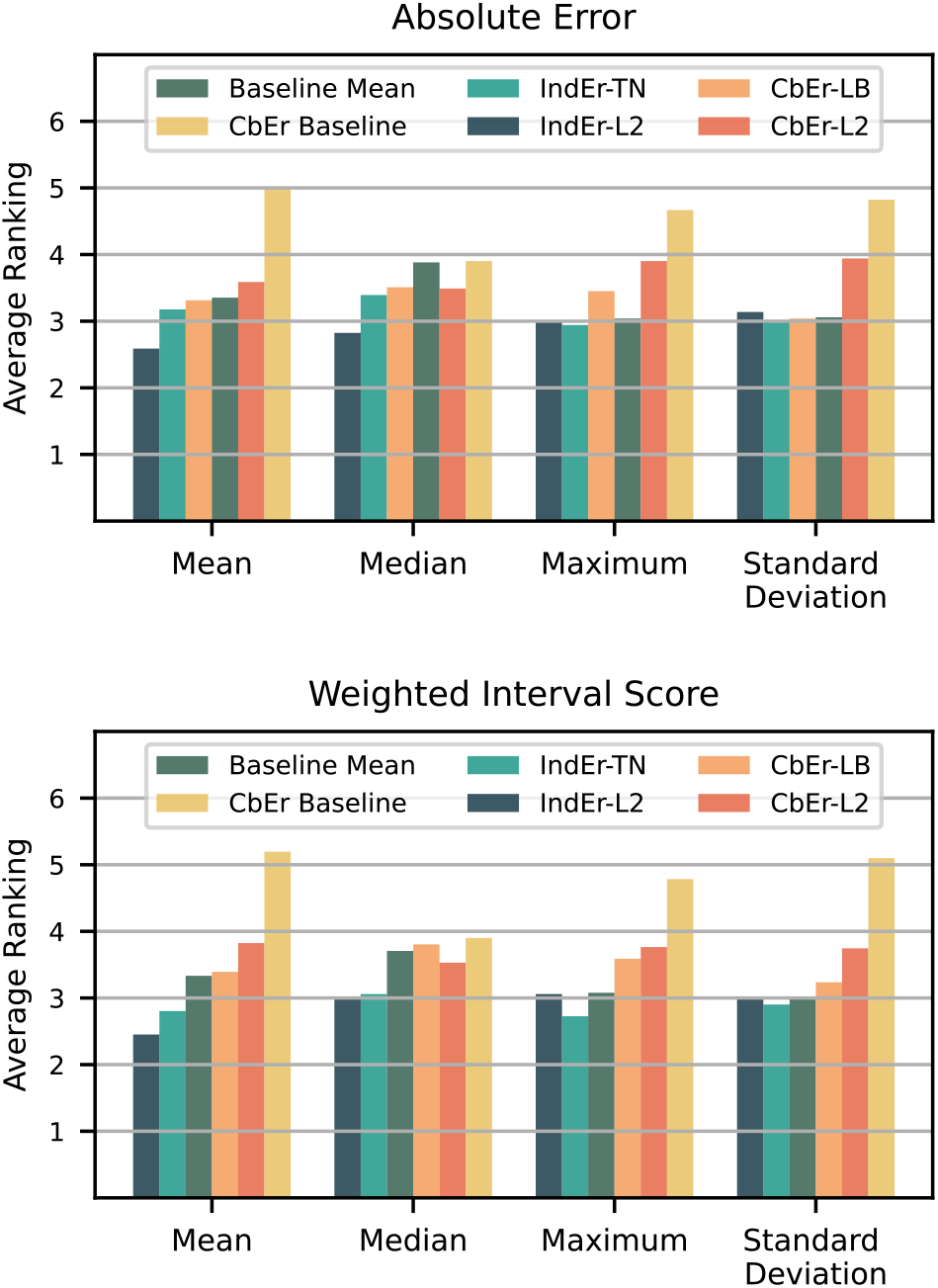
Average rankings of ensemble model mean, median, maximum, and standard deviation of the absolute error and weighted interval score in each state, where a smaller rank indicates smaller error compared to other methods.

**Table 1.** Average of model mean, median, and maximum absolute error in each state, sorted by the average mean absolute error. An outlier (Ohio) was manually removed before calculating the mean over the states.

|  | Mean | Median | Maximum |
| --- | --- | --- | --- |
| IndEr-L2 | 38.21 | 26.71 | 143.86 |
| IndEr-TN | 39.04 | 27.61 | 143.76 |
| Baseline Mean | 39.60 | 28.80 | 150.77 |
| CbEr-LB | 39.63 | 28.91 | 150.53 |
| CbEr-L2 | 39.74 | 27.72 | 156.91 |
| CbEr Baseline | 42.38 | 29.22 | 168.77 |

From examining the absolute error rankings and mean statistics, Table 1 and Figure 2, a few trends become clear. The two Individual Error (IndEr) methods had smaller aggregated error statistics than the Combined Error (CbEr) methods. Second, the CbEr Baseline method displayed poorer performance than the Baseline Mean and all other methods on average, and was the only model with average rankings above four, where the maximum rank was six. The weighted interval score rankings displayed similar trends compared to the absolute error rankings.

The IndEr-L2 method was the top performer for mean and median absolute error and WIS across states, but it is worth noting that the IndEr-TN method and Baseline Mean provided competitive rankings among the maximum absolute error, with the IndEr-TN method having the smallest average ranking. For example, in Figure 3, one can see how for the ensembles of New Jersey forecast models, the maximum IndEr-L2 absolute error was larger than that of the IndEr-TN method, thus would be ranked lower for that state. However, the IndEr-L2 errors are smaller on average across time, with a MAE of 23.51 compared to 27.98. The mean values in Table 1 show that the Baseline Mean’s maximums may be larger in magnitude, which is also demonstrated in Figure 3. Thus, the similar absolute error rankings of the IndEr-TN and Baseline Mean compared to the IndEr-L2 method may be insufficient to offset the method’s better performance on the other metrics, such as the mean absolute error.

**Figure 3.**
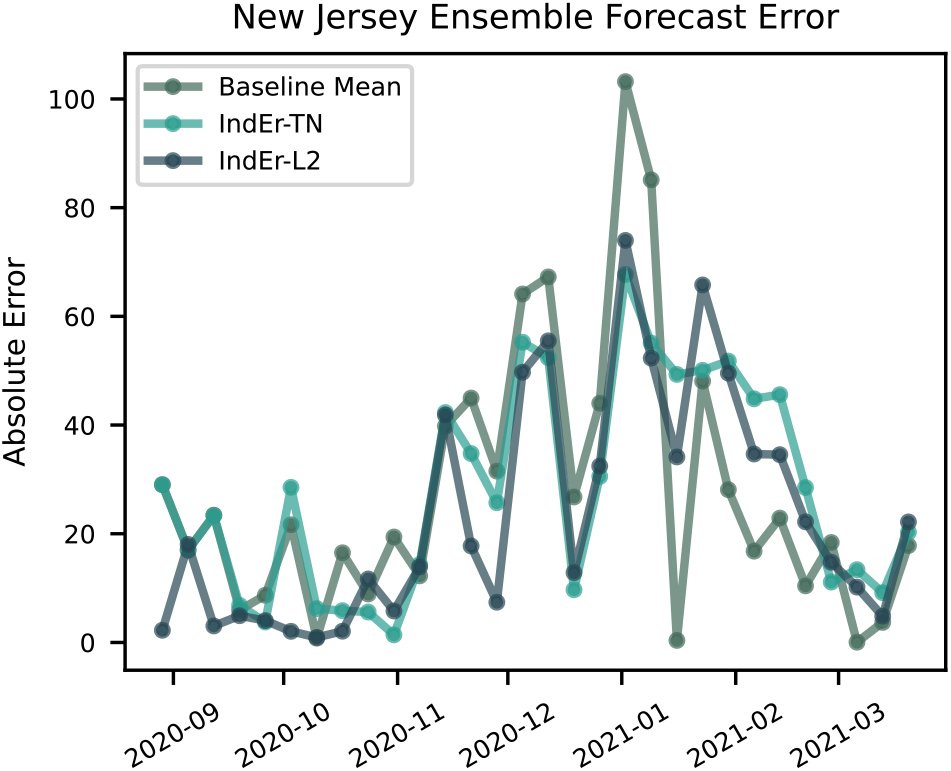
Absolute error of Individual Error (IndEr) ensemble forecasts of New Jersey 1-week-ahead incident deaths. This is an example of the 30 weeks of forecast error calculated for each state and ensemble method.

The MAE of the two methods that use L2 regularization for different parameter values in Figure 1 show a notable trend. When the hyperparameter *α* is increased from 0 to 0.02, the error drops drastically for the IndEr-L2 method. This indicates that choosing the single best model during the training period (*α* = 0), as one would in typical cross-validation, resulted in significantly more error than when even a slight encouragement to include more models is added. The shape of the two curves indicate that not all models should be included in an ensemble: the error increases as *α* increases and as weights approach equality. Additionally, the comparison of the IndEr-L2 method and CbEr-L2 method across the same spectrum of regularization parameters shows that for the US 1-week-ahead incident death forecasts, the IndEr-L2 ensemble produced forecasts with lower errors for all *α* values greater than 0.

## 4 Discussion

Multiple ensemble methods were implemented on publicly available COVID-19 forecasts of 1-week-ahead incident deaths and were evaluated across time and location. We created a new forecast ensemble method that we refer to as Individual Error with L2 Regularization or IndEr-L2, in which the weighted sum of component model point forecast root-mean-squared error (RMSE) is minimized with L2 regularization. This method demonstrated improved performance on the test set relative to other methods including a baseline mean ensemble, a method where the top performing models were selected with equal weight, and variations of a typical stacking ensemble algorithm where the error of the combined models was minimized. The IndEr-L2 ensemble captures the varying performance of models across location targets, unlike other COVID-19 ensemble methods that do not take performance into account (11). While this method is composed of well-known statistics, the particular combination of the two components of the objective function and its application to disease forecasting is new. Additionally, the simplicity of this ensemble method compared to techniques such as Bayesian Model Averaging (BMA) is an advantage because it lowers the threshold of understanding the results, and may thus increase trust from stakeholders. Though all ensembles were trained on only point forecasts, the respective performance of the IndEr-L2 method for quantile ensembles was similar to that of the point ensembles, demonstrating the efficacy of training an ensemble in this manner.

The success of the IndEr-L2 approach relative to the methods that optimize over the error of the combined models, Combined Error or CbEr methods, can be explained by considering the challenges of predicting COVID-19 outcomes. Forecasters have needed to continuously change their underlying assumptions and inputs throughout the pandemic to account for rapidly changing conditions (e.g., new variants of the disease, the introduction of vaccines). Compared to similar ensemble work in which influenza forecasts were retrospectively generated for previous seasons using stable modeling frameworks (7), the adjustment of parameters during the COVID-19 season may have led to an unpredictable relationship between the combined models and the observed data. This is the essence of what stacked ensembles are meant to learn (14). Therefore, if this learned relationship was subject to change from week to week, the CbEr ensembles may have forecasted poorly. This was seen especially in the results of the CbEr Baseline, which was un-regularized and often performed worse than simply averaging all valid models. In contrast, the IndEr methods simply measured individual model error, or rather the relationship between each model and the observed data, and the successful results demonstrated that this relationship was better maintained across time than that of the combination of models and observed data.

Beyond the immediate results, the IndEr-L2 method also has an advantage of extensibility. The function is a sum of sums, thus it is a convex function of its parameters, the weights. One could easily replace the RMSE with the average weighted interval score or the maximum absolute error. The same cannot be said for the CbEr methods; if the weighted interval score were to be used, the objective function would become non-differentiable and potentially non-convex if multimodal models are present. This would require a more advanced solver and increased computational resources and run times.

The most notable future extension of this study is refining the method of hyperparameter selection. Ensemble results from half of the dates in the test set for the US and select states were used to choose hyperparameters for each method, which is an in-sample selection. To fully assess real-time performance of the ensembles, hyperparameter selection could be performed on a historical ensemble forecast test set. We note this would be difficult when little data is available in the early stages of the pandemic, especially when historical seasons are lacking. Previous research was able to leverage past seasons of influenza forecasts to optimize parameters (7). Research into hyperparameter and overall ensemble method selection during the early stages of a new disease outbreak is warranted. In the later stages, refinements could be made to find more specific hyperparameter values. Performing global optimizations separately for different locations and allowing a new selection for each week could improve performance, though caution will need to be exercised to prevent overfitting.

Future work that could address the limitations of this research and provide further confidence in the results include applying methods to additional forecast targets, and performing sensitivity analysis on the parameters and methods. Forecasting two-to-four weeks ahead and modifying the outcome to incident cases consistently results in more intra-model uncertainty, inter-model variation, and poorer performance on empirical data than the 1-week-ahead death forecasts studied. Applying the methodologies described here to other outcomes and further time horizons would enable a more comprehensive assessment of the ensemble methods. When forecast data are limited, especially early in an epidemic, a small number of model forecasts could have a large influence on ensemble method performance. Repeated calculations of metrics with data re-sampling would add certainty to the comparison of ensemble method performances.

The value of performance-based ensemble models in disease forecasting was demonstrated in this study. We saw that combining more than one model resulted in smaller error than choosing the single best model based on performance of US incident death forecasts. Additionally, using historical performance to inform an unequal-weighted sum ensemble demonstrated improved error when compared to a simple average of all models across US states. Minimizing a distanced-based measure of combined model performance in the training phase is not recommended for a rapidly-evolving situation like COVID-19.Instead, optimization of a weighted sum of individual model error with regularization to push weights towards equality should be considered. Implementing the outlined future work could provide additional confidence in the aforementioned method.

## Data Availability

All data produced in the present study are available upon reasonable request to the authors

https://covid19forecasthub.org/

## Acknowledgements

The authors thank Christopher Ratto and Shelby Wilson, Johns Hopkins University Applied Physics Laboratory (JHU/APL), who provided technical manuscript reviews. We also thank Mark J. Panaggio, JHU/APL, for providing helpful code for the interval score implementation, and Perry Wilson, JHU/APL, for informative conversations on databases. Declarations of interest: none.

